# Regional Variation in Antenatal Late Preterm Steroid Use following the ALPS Trial

**DOI:** 10.1101/2023.05.25.23290522

**Authors:** Taylor S. Freret, Jessica L. Cohen, Cynthia Gyamfi-Bannerman, Anjali J. Kaimal, Scott A Lorch, Jason D. Wright, Alexander Melamed, Mark A. Clapp

## Abstract

**Objective:** To assess regional variation in the use of late preterm steroids use after the publication of the Antenatal Late Preterm Steroids (ALPS) Trial and to understand factors associated with a region’s pace of adoption.

**Methods:** This was a repeated cross-sectional study using US natality data across hospital referral regions (HRRs) within the US from February 2015 to October 2017. Inclusion criteria included live-born, non-anomalous, singleton, late preterm (34-36 completed weeks of gestation) neonates born to individuals without pregestational diabetes. HRRs were categorized as either a “slower” or “faster” adopter of antenatal late preterm steroids based on the observed versus expected pace of antenatal steroid adoption in a 1-year period after the trial’s dissemination compared to a 1-year pre-dissemination period. Patient and regional factors hypothesized *a priori* to be associated with the uptake of late preterm steroids were compared between faster and slower adopters.

**Results:** There were 666,097 late preterm births in 282 HRRs during the study period. Of the included HRRs, 136 (48.2%) were considered faster adopters and 146 (51.8%) slower adopters. Faster adopters increased their steroid use by 12.1-percentage points (5.9% to 18.0%) compared to a 5.5-point increase (3.7% to 9.2%) among slower adopters (p<0.001). Most examined patient and regional factors were not associated with a region’s pace of adoption. The only factor associated with being a faster adopter was the regional prevalence of prior preterm birth (adjusted odds ratio 2.04, 95% confidence interval 1.48-2.82).

**Conclusion:** There was widespread geographic variation in the adoption of antenatal steroid administration for late preterm births that largely remained unexplained by population factors. Future research should focus on patient, provider, and system-level factors that may be influencing the adoption of late preterm steroids. Namely, this work can provide insights to barriers to timely or equitable access to new evidence-based practices and can guide future dissemination strategies with the goal of more uniform adoption.

## Introduction

The publication of the Antenatal Late Preterm Steroids (ALPS) trial in February 2016 demonstrated that antenatal administration of betamethasone in the late preterm period (between 34-36 weeks of gestation) for individuals at high risk of delivery decreased neonatal respiratory morbidity.^1^ Following its publication, steroid exposure among infants born in the late preterm period increased by more than 200% within months nationally.^2-5^ This practice change was rapid in comparison to prior studies examining the pace of evidence dissemination in clinical medicine.^6^ However, steroid exposure also increased among groups not included in the original trial, including individuals with pregestational diabetes and multiple gestation pregnancies, where the benefit of late preterm steroids is unknown, and among term infants, in which there is no hypothesized benefit.^7, 8^ These findings emphasize the need for more evidence regarding how and why practice changed quickly so as to better inform policies and care practices around the appropriate and timely use of this intervention.

The primary objective for this study was to examine patterns of regional variation in the administration of antenatal late preterm steroids and determine if regional factors may be associated with the pace of adoption of this practice. Secondarily, we aimed to quantify the proportion of regional variation that was explained by patient and regional factors. As substantial geographic variation exists in the provision of other healthcare services, we hypothesized that there would be measurable variation in late preterm steroid use across hospital referral regions (HRR) in the US and that variation would be attributed to differences in patient case mix.^9-12^

## Methods

### Study Details

This repeated cross-sectional study was conducted from November 15, 2022, to January 13, 2023, using county-identified data that were obtained with permission from the National Center for Health Statistics.^13^ The project was classified as non-human subjects research by the Mass General Brigham Human Subjects Research Committee and was therefore exempt from institutional review board approval. We followed guidelines from the Strengthening the Reporting of Observational Studies in Epidemiology statement.^14^

Analyses were performed using Stata MP software, version 15.1 (StataCorp LLC). Due to the number of comparators and the large sample size, an *a priori* 2-sided p-value of <0.001 was considered statistically significant.

### ALPS Trial Dissemination and Analysis Periods

The ALPS trial was first available online in February 2016, published in print in April 2016, and included in updated clinical guidance in August and October 2016 from the Society for Maternal-Fetal Medicine (SMFM) and the American College of Obstetricians and Gynecologists (ACOG), respectively.^1, 3-5^ Thus, we considered February to October 2016 to be the period of evidence dissemination, as designated in previous work.^2^ Data from 12-month observational period before (February 2015 to January 2016, “pre-period”) and after (November 2016 to October 2017, “post-period”) the dissemination period were used to assess variation in late preterm steroid use and the pace of adoption.

### Unit of Analysis

Because of the small number of preterm births in many counties, geographic analysis was performed at the hospital referral region (HRR) level. HRRs were defined by the Dartmouth Atlas Project and represent regional health care markets for tertiary medical care, and by definition include at least one hospital where both major cardiovascular and neurosurgical procedures are performed.^15^ County-level federal information processing system (FIPS) codes from delivery locations were mapped to hospital referral regions using previously published crosswalks.^15^

To facilitate meaningful comparisons across the time periods and reduce noise in the estimates, two HRR inclusion criteria were set: 1) more than 100 eligible late preterm births in the pre-period, and 2) steroid exposure data available for all eligible births. Eligible births were defined as liveborn, non-anomalous singleton neonates born between 34 and 36 completed weeks of gestation to individuals without pregestational diabetes (i.e., participants most similar to those in the ALPS trial).^2^ Births were not eligible for inclusion if antenatal steroid exposure was unknown, if data elements were not reliably reported per the Natality Data User Guides, or if the FIPS code for the location in which they delivered was unknown.^16^

### Defining Pace of Adoption

The primary outcome of interest was HRR-level adoption status of late preterm use, designated as either “slower” or “faster” adopters based on the extent to which observed rates of late preterm births exposed to antenatal steroids compared to the expected rates in the 12-month period following the ALPS trial dissemination. Expected rates were derived from a patient-level logistic regression model that included patient factors that were hypothesized *a priori* to influence adoption: maternal age (continuous); parity (categorized as nulliparous or multiparous); race (pre-categorized in the US natality data as White, Black, American Indian/Alaskan Native, or Asian or Pacific Islander); ethnicity (pre-categorized as Hispanic or non-Hispanic in the US natality data); primary payer for delivery (categorized as private, public, self-pay, or other); and education (binary variable indicating some college education); maternal comorbidities (binary variables indicating the presence or absence of pregnancy-related and pre-existing hypertensive disorders and gestational diabetes); week of gestation at delivery (categorical); an indicator if a maternal transfer occurred before delivery; and an indicator if the patient delivered in the same county as their residential address. Race and ethnicity were included not based on a biological hypothesis but because preexisting literature has suggested rates of steroid administration vary across these groups.^17^ As an underlying increasing trend in antenatal steroid exposure was observed in the pre-period, which is hypothesized to local changes in reporting practices, the regression model also adjusted for pre-period steroid exposure trends in each HRR (determined by including a categorical variable for each HRR and an interaction term between HRR and month in the pre-period in the model).

Using this regression model, a predicted probability for steroid administration in the post-period was generated for each eligible patient. These predicted probabilities were summed within each HRR to determine the expected number of patients who received steroids and the steroid administration rate in the post-period. The difference in the observed and expected rates of steroid exposure in the post-period was calculated for each HRR. Rate differences less than the median were considered “slower” adopters; HRRs with rate differences equal to or greater than the median were considered “faster” adopters.

### Factors Associated with Adopter Status

Once adopter status had been assigned, we examined for regional factors that were associated with adoption pace. The following HRR population characteristics were compared between slower and faster adopters at the HRR level in the pre-period: mean maternal age, race (White, Black, Native Hawaiian or Pacific Islander, or Asian, per pre-defined US natality data categories), Hispanic ethnicity, education, gestational diabetes, gestational hypertension, chronic hypertension, prior preterm birth, and primary payer for delivery admission. Except for maternal age (which used population mean), all variables were expressed as a percentage of late preterm births. Similarly, the following HRR regional characteristics were compared between slower and faster adopters in the pre-period: delivery provider (percentage of late preterm births attend by a physician, midwife, or other), rate of infant transfers, number of hospitals with obstetric care per square mile, number of births per obstetric bed, number of births per higher-level pediatric bed (neonatal intensive care unit or special care nursery), population density, and total geographic area (sq. mi). Variables related to the number of hospitals, beds, births, or geographic area were obtained from the 2017 Area Health Resource Files, which were publicly available from the US Health Resources and Services Administration.^18^ These covariates were selected *a priori* based on known associations with preterm birth or hypothesized to be related to an area’s tendency to adopt late preterm steroids into practice.^19-22^ Comparisons were made using Student’s t-test or Wilcoxon rank-sum test, as appropriate. Absolute standardized mean differences between the groups were also calculated for comparison.

Then, a multivariable logistic regression was constructed using the covariates listed above to identify factors associated with adopter status in the post-period. To facilitate the interpretation of the coefficients, independent variables that represented population percentages were standardized before regression analysis and variables involving bed or population density were scaled based on birth volumes.

### Quantifying Regional Variation

To quantify the proportion of variation that could be attributed to regional differences in steroid use (i.e., inter-regional practice variation), a series of patient-level hierarchical random-effects linear regression models were constructed where the outcome was steroid exposure among late preterm births. The degree of inter-regional practice variation, estimated via the intraclass correlation coefficient, was determined using methods that have been previously described for similar study questions.^23, 24^ The series included the following models: 1) random effect of the HRR only (base model); 2) HRR random-effect and known regional characteristics; and 3) HRR random-effect term, known regional characteristics, and patient factors (i.e., case mix adjustment). The regional factors included the slope and intercept of steroid use in HRR from pre-period (to adjust for pre-existing trends in use), the number of OB hospitals per total geographic area, total births per obstetric beds, total births per higher-level pediatric care beds, population density, and total geographic area. The patient factors included: maternal age, race, Hispanic ethnicity, gestational age (categorized as completed weeks of gestation), gestational diabetes, pre-existing and pregnancy-related hypertension, provider type, delivery payer type, maternal education status, prior preterm birth, and whether the patient was transferred.

## Results

There are 306 HRRs in the United States; 24 were excluded as they had fewer than 100 births recorded in the pre-period. From the 282 included HRRs, a total of 666,097 births occurred between February 1, 2015, and October 31, 2017: 239,372 (35.9%) occurring in the pre-period, 182,352 (27.4%) in the dissemination period, and 244,373 (36.7%) occurring in the post-period (Figure 1).

**Figure 1:**
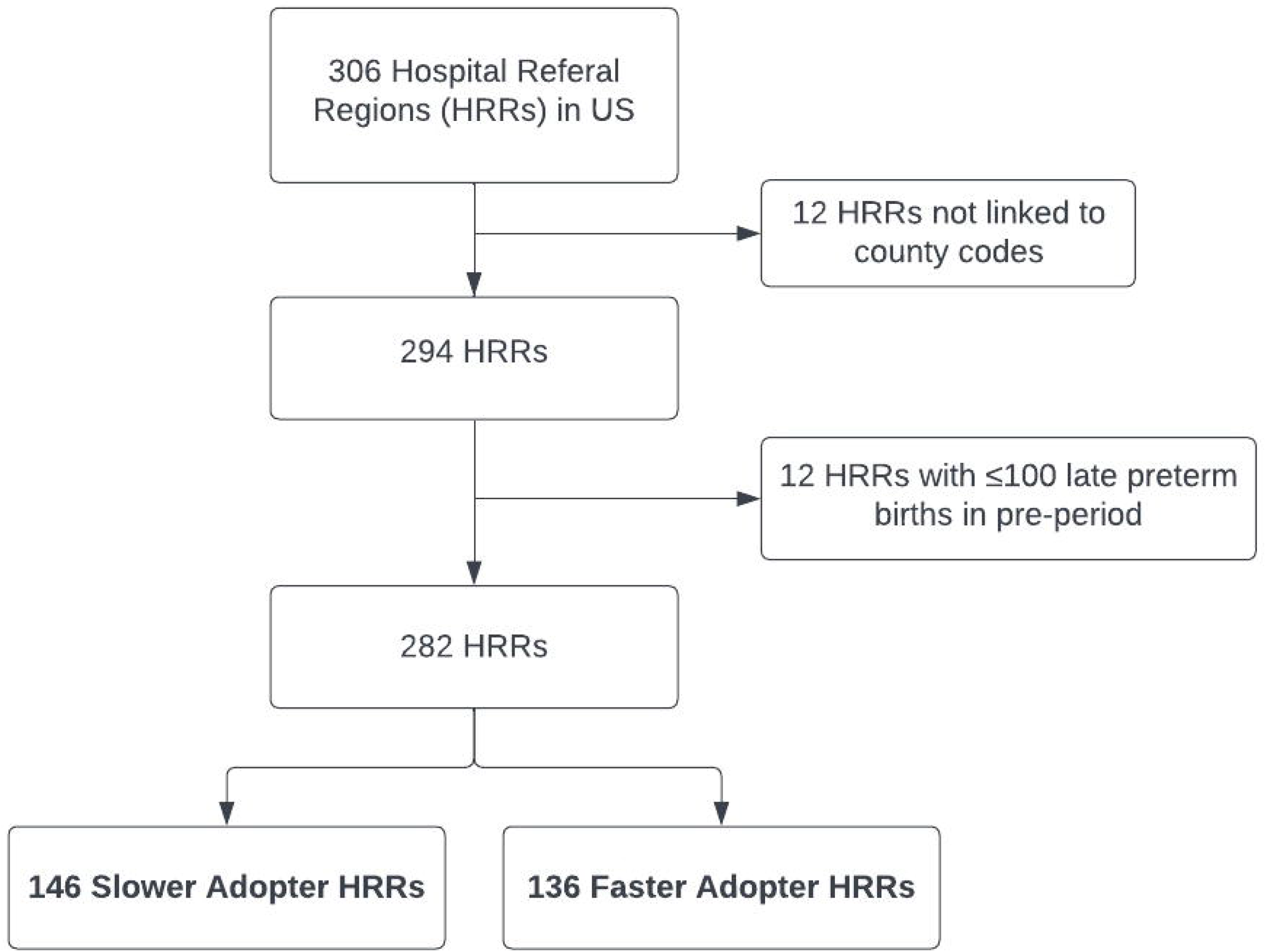
Inclusion diagram Adopter status (slower or faster adopter) determined by the difference between observed and expected steroid use in the year following the Antenatal Late Preterm Steroid (ALPS) Trial relative to the median change in steroid use among all hospital referral regions.

After the publication of the ALPS Trial, late preterm steroid use among HRRs ranged from 0 to 47.4% with median 11.7% (IQR 6.85-18.6%). There was widespread geographic variation noted relative to the expected steroid rate in the post period, with differences in the observed and expected rates ranging from 94.8% less than expected to 33.8% more than expected based on patient characteristics and pre-period trends (median 4.9% more than expected (IQR -0.77, 11.0%)) (Figure 2). Of the 282 HRRs, 136 (48.2%) were designated as faster adopters and 146 (51.8%) as slower adopters. In HRRs categorized as faster adopters, steroid use increased from 5.9% to 18.0% between the pre- and post-periods. In comparison, in slower adopting HRRs, steroid use increased from 3.7% to 9.2% over the same period. This corresponded to a 12.1 percentage point increase in steroid-exposed births after the dissemination period in HRRs designated as faster adopters compared to a 5.5-point increase among slower adopters (p<0.001) (Figure 3).

**Figure 2:**
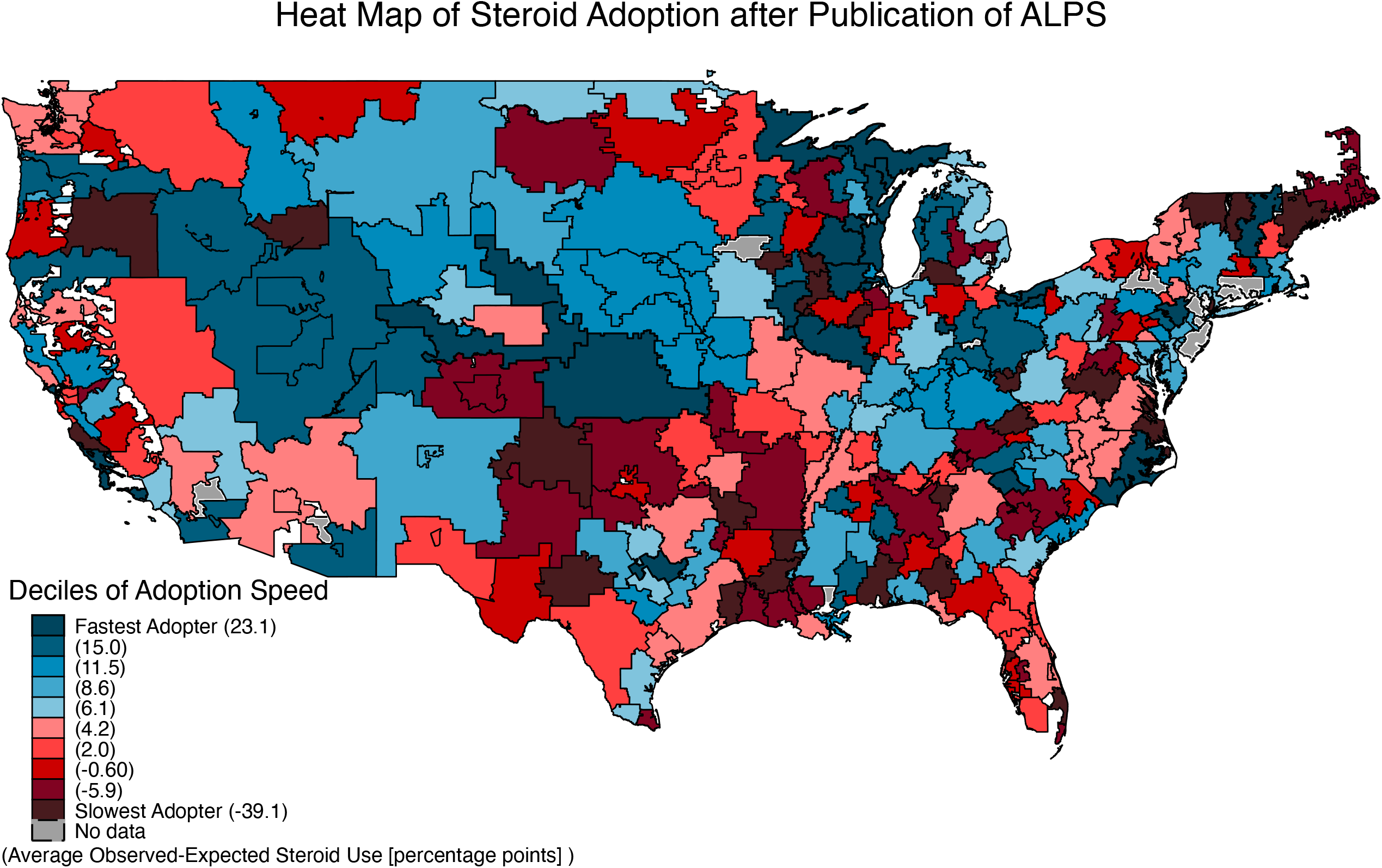
Heat map of late preterm steroid adoption after the ALPS Trial by hospital referral region (HRR) Adopter status determined by the difference between observed and expected steroid use in the post-period relative to the median change among all hospital referral regions in the year following the Antenatal Late Preterm Steroid (ALPS) Trial.

**Figure 3:**
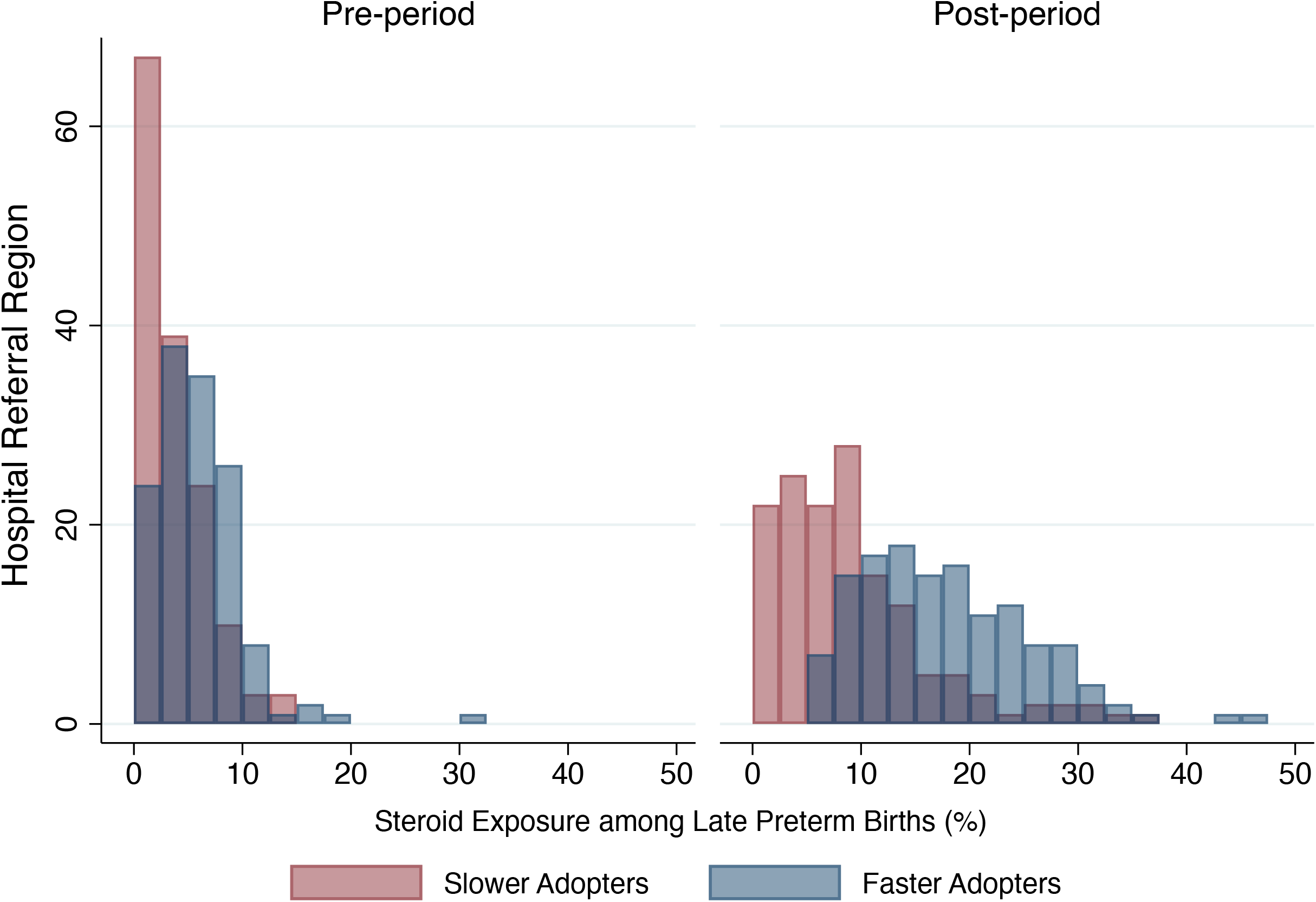
Histogram of antenatal late preterm steroid use in the pre- and post-periods by hospital referral region adopter status In the year before the Antenatal Late Preterm Steroid (ALPS) Trial (pre-period), there was similar late preterm steroid use between the slower and faster adopter hospital referral regions. In the year following the trial (post-period), the use diverged between the two groups.

Baseline characteristics between faster and slower adopters were similar in the pre-period (Table 1). Two characteristics were statistically different: faster adopters had a lower proportion of patients with some college experience (45.9 vs 49.4%, p<0.001) and a higher rate of patients with a prior preterm birth (9.4 vs 7.2%, p<0.001) compared to slower adopter regions.

**Table 1:** Baseline characteristics of hospital referral regions (HRR) in pre-period

|  | **Fast Adopters (n=136)** | **Slow Adopters (n=146)** | **Absolute Standard Mean Difference** |
| --- | --- | --- | --- |
| **Population Characteristics** |  |  |  |
| Maternal age | 28.15 (1.26) | 27.73 (1.16) | 0.31 |
| Race<sup>†</sup> |  |  |  |
| White | 75.88 (15.53) | 75.02 (15.71) | 0.05 |
| Black | 16.79 (16.03) | 18.43 (15.70) | 0.10 |
| Asian or Pacific Islander | 5.07 (5.79) | 4.71 (7.59) | 0.06 |
| Native Hawaiian/Alaskan Native | 2.26 (5.41) | 1.84 (4.25) | 0.10 |
| Hispanic Ethnicity<sup>†</sup> | 18.80 (17.30) | 19.35 (19.64) | 0.03 |
| Some College Education<sup>†</sup> | 45.90 (8.04)* | 49.39 (9.21)* | 0.39 |
| Gestational Diabetes<sup>†</sup> | 8.34 (2.79) | 7.56 (2.61) | 0.30 |
| Gestational Hypertension<sup>†</sup> | 11.70 (3.78) | 10.58 (3.39) | 0.25 |
| Chronic Hypertension<sup>†</sup> | 3.17 (1.94) | 2.79 (1.63) | 0.21 |
| Prior Preterm Birth<sup>†</sup> | 9.40 (4.17)* | 7.18 (3.88)* | 0.58 |
| Insurance<sup>†</sup> |  |  |  |
| Private | 43.13 (11.80) | 38.76 (12.31) | 0.33 |
| Public | 49.70 (11.69) | 53.47 (12.57) | 0.27 |
| Self-Pay | 3.20 (2.88) | 3.56 (3.35) | 0.10 |
| Other | 3.96 (5.14) | 4.22 (6.10) | 0.07 |
| **Health System Characteristics** |  |  |  |
| Gestational Age at Delivery<sup>†</sup> |  |  |  |
| 34 weeks | 17.92 (2.58) | 17.51 (2.65) | 0.06 |
| 35 weeks | 28.00 (2.77) | 28.53 (2.38) | 0.22 |
| 36 weeks | 54.08 (3.81) | 53.96 (3.68) | 0.10 |
| Delivery Provider<sup>†</sup> |  |  |  |
| MD/DO | 92.03 (6.90) | 91.76 (6.49) | 0.02 |
| CNM | 6.95 (6.35) | 7.11 (6.20) | 0.01 |
| Other | 1.02 (1.34) | 1.14 (1.18) | 0.14 |
| Infant Transfer<sup>†</sup> | 3.08 (3.96) | 3.04 (3.71) | 0.02 |
| OB Hospital Density (per 100 sq. mi) | 0.71 (0.37, 1.71) | 0.57 (0.26, 1.13) | 0.28 |
| Total Births per OB Bed | 70.1 (58.8, 87.6) | 70.8 (56.5, 91.8) | 0.07 |
| Total Births per Higher-level Pediatric Bed | 145.9 (117.6, 202.1) | 155.0 (113.0, 212.1) | 0.19 |
| **Geographic Characteristics** |  |  |  |
| Population Density (per sq. mi) | 344.9 (180.7, 936.3) | 330.5 (144.8, 687.3) | 0.26 |
| Total Area (sq. mi) | 760.1 (535.8, 1264.8) | 863.0 (573.5, 1423.8) | 0.05 |

Univariate statistics listed as mean (standard deviation) or median (interquartile range). \*P-value for the comparison trend <0.001, considered the threshold for significance. †Listed as percent of HRR population. Obstetric (OB) hospital density refers to the number of hospitals with obstetric care, as defined by the 2017 Area Health Resources File per total HRR land area. Total births refer to the total number of births in the HRR (including all preterm and term births). Higher-level pediatric beds includes both neonatal intensive care unit and special care nursery beds.

**Table 2:** Multivariable Predictors of HRR Late Preterm Steroid Adopter Status.

| **Population Characteristics** | **Adjusted Odds Ratio** | **p-value** |
| --- | --- | --- |
| Maternal age | 0.95 [0.60 - 1.52] | 0.83 |
| Race<sup>†</sup> |  |  |
| White | Referent |  |
| Non-White | 0.95 [0.65 - 1.38] | 0.78 |
| Hispanic Ethnicity<sup>†</sup> | 1.12 [0.77 - 1.65] | 0.55 |
| Some College Education<sup>†</sup> | 0.74 [0.47 - 1.16] | 0.19 |
| Gestational Diabetes<sup>†</sup> | 1.01 [0.72 - 1.41] | 0.96 |
| Gestational Hypertension<sup>†</sup> | 1.14 [0.84 - 1.53] | 0.40 |
| Chronic Hypertension<sup>†</sup> | 1.32 [0.93 - 1.86] | 0.12 |
| *Prior Preterm Birth*<sup>†</sup> | *2.04 [1.48 - 2.82]* | *<0.001* |
| Insurance<sup>†</sup> |  | 0.62 |
| Private | Referent |  |
| Non-Private | 1.13 [0.69 - 1.86] |  |
| **Health System Characteristics<sup>†</sup>** |  |  |
| Gestational Age at Delivery<sup>†</sup> |  |  |
| 34-35 | 0.68 [0.47 - 0.99] | 0.04 |
| 36 | Referent |  |
| Delivery Provider<sup>†</sup> |  |  |
| Doctor | Referent |  |
| Midwife/Other | 1.09 [0.83 - 1.44] | 0.53 |
| Infant Transfer<sup>†</sup> | 0.87 [0.62 - 1.20] | 0.39 |
| OB Hospital Density (per 100 sq. mi) | 1.10 [0.84 - 1.44] | 0.49 |
| Total Births (Hundreds) per OB Bed | 1.67 [0.65 - 4.32] | 0.29 |
| Total Births (Hundreds) per Higher-level Pediatric Bed | 0.88 [0.73 - 1.07] | 0.21 |
| **Geographic Characteristics** |  |  |
| Population Density (in 100s per sq. mi) | 1.04 [0.99 - 1.10] | 0.08 |
| Total Area (100 sq. mi) | 1.00 [0.98 - 1.02] | 0.76 |

Regression predicting faster adopter status includes all variables listed.
<sup>†</sup> As the analysis was performed at regional (not patient) level, characteristics are expressed as percent of the population. When the underlying patient-level variable was categorical (race, insurance, gestational at delivery, delivery provider), the most frequent response (i.e., population majority) was compared to the other groups combined in this regional-level analysis. We acknowledge a limitation of this approach is that it precludes a more granular assessment of the grouped categories and assumes that the grouped categories have a similar relationship in relation to the comparator.

In the multivariable logistic regression analysis, the percent of the population with a prior preterm birth was the only factor significantly associated with faster adopter status (adjusted odds ratio (aOR) 2.04, 95% confidence interval (CI) 1.48-2.82). This coefficient can be interpreted as the change in odds associated with a one-standard deviation increase in the population rate of prior preterm birth. No other observed regional factors were significantly related to the pace of adoption.

When quantified using the random-effects models, there was a large amount of measurable variation in steroid exposure among late preterm births attributable to practice differences between HRRs (19.9%, 95% CI 17.1-23.1%). When adjusting for observed regional characteristics, the proportion of variation attributed to inter-regional practice differences decreased to 10.9% (95% CI 9.0-13.0%). However, it was largely unaffected by patient case-mix adjustment in the final model (10.7%, 95% CI 8.8-12.8%).

## Discussion

The ALPS Trial demonstrated that antenatal steroid administration during the late preterm period reduced neonatal respiratory morbidity, a major cause of healthcare resource utilization in the US, with a secondary analysis demonstrating that late preterm steroids are likely cost-effective.^25^ These findings have also been shown outside of a clinical trial setting in a subsequent quasi-experimental study.^1, 2^ Professional societies in the United States (namely, ACOG and SMFM) have recommended the use of late preterm steroids in eligible populations. Despite these recommendations, we observed significant regional variation in the adoption of this practice across the US in addition to overall high levels of non-adoption. More than 10% of the variation between regional steroid use remained unexplained after accounting for regional factors, such as hospital density or bed availability, that might be expected to influence adoption. Moreover, essentially no additional variation was explained by accounting for patient factors known to be associated with preterm delivery, including maternal hypertension and diabetes. This non-uniform uptake raises additional questions as to why this multi-center randomized trial and nationwide dissemination strategy did not result in more uniform or predictable widespread uptake.

Only one regional factor was associated with ALPS adoption: the population prevalence of prior preterm birth. The ALPS trial highlighted the challenge of appropriately identifying who is at risk for preterm birth, with nearly 20% of the trial subject’s ultimately delivering at full term.^1^ As prior preterm birth is a strong predictor for recurrent preterm birth, we hypothesize that in areas where the *a priori* risk of preterm birth may be higher (e.g., more individuals with a prior preterm birth), clinicians may be more likely to recommend and administer late preterm steroids and that health systems may be better equipped to administer late preterm steroids due to increased underlying need. In contrast, the population prevalence of hypertensive disorders of pregnancy, a common medical indication for a late preterm birth, was not associated with ALPS adoption, although the ascertainment of the timing and severity of this condition is not well known from the natality data and should continue to be explored.

As no other regional characteristics (including region size, population density, obstetric and neonatal bed availability) were associated with adoption, late preterm steroid use may be influenced by more granular provider- and hospital-level practices and patient preferences, many of which cannot be measured in population-based data sources. Future quantitative and qualitative work should seek to identify more local sources of practice variation and identify barriers to adoption. These results will inform policies to guide timely and appropriate use of late preterm steroids and inform the dissemination of new evidence-based medicine practices in obstetrics.

The strengths of this study include the use of US natality data, which represents a complete sample of late preterm births in the US, and methodology incorporating the use of pre-existing time trends (in addition to fixed factors) in each HRR to predict expected steroid exposure in the post-period. The natality data is one of the only sources where antenatal steroid exposure, birth outcomes, and maternal information can be all ascertained.

There are also several limitations to our study. HRRs were not constructed around perinatal or obstetric care services specifically, though they are commonly used to examine regional practice variation in other areas of medicine. As each HRR encompasses many counties, the case-mix, health systems, and geographic characteristics represent averages that are expected to trend toward the population mean, which may limit the ability to discern the impact of any specific characteristic. Additionally, we were not able to control for some factors that may have influenced the clinical or shared decision-making around steroid administration, such as indication or likelihood for a late preterm delivery. The exact timing of steroid administration is unknown in the natality data.

Certain elements of birth certificate data may be subject to low sensitivity or specificity, including steroid administration; it is likely that steroid administration rates are underreported but not systematically in a way that negates the study findings.^26^ We accounted for potentially inaccurate reporting by excluding births who had concerns regarding data reliability per the yearly Natality Data User Guides. It is also possible that birth certificate reporting practices varied between each HRR over time, in which case the relative changes of the observed and expected rates of steroid exposure may not have been uniform. We accounted for this potential variation by adjusting the models for pre-existing trends in steroid use each HRR; however, we are not able to predict or observe which HRRs may have differentially increased reporting after the ALPS Trial publication.

There was widespread regional variation in the use of late preterm steroids after publication of the ALPS Trial. After accounting for patient and regional factors, a significant amount of inter-regional variation remained unexplained, suggesting that the evidence was not adopted uniformly nor predictably. This unexplained variation remains a high priority for future research, including studies examining variation at a more granular level (e.g., patient-, clinician- and hospital-level factors), to understand why the adoption of late preterm steroids was so varied. Namely, it can provide insights as to why some patients may or may not have timely or equitable access to new evidence-based practices and guide future dissemination strategies with the goal of more uniform adoption.

## Data Availability

Data used to construct this analysis can be found at: https://www.cdc.gov/nchs/data_access/vitalstatsonline.htm

## Acknowledgments

The authors have no acknowledgments.

